# Executive functions in children and adolescents with ADHD, ASD, and Affective Disorders: a differential exploratory analysis

**DOI:** 10.1101/2025.06.17.25329783

**Authors:** Tiago Figueiredo, Leandro Malloy-Diniz

## Abstract

Executive function cognitive deficits and behavioral issues are implicated in neurodevelopmental disorders such as Autism Spectrum Disorder (ASD), Attention Deficit/Hyperactivity Disorder (ADHD), and affective disorders. The current study enrolled 134 children and adolescents (aged 6-17) who were referred to a specialized service. Participants were categorized into groups based on ADHD, ASD, and affective disorders to conduct an exploratory analysis of multilevel executive dysfunctions. Guardians completed the Strengths and Difficulties Questionnaire (SDQ) and the Behavior Rating Inventory of Executive Function-Second Edition (BRIEF-2) as part of the assessment process. Children engaged in tasks designed to evaluate both affective and non-affective executive functions. Dimensional exploratory analyses revealed that the ADHD and ASD groups exhibited greater cognitive non-affective executive deficits. However, performance non affective executive functions did not significantly differentiate participants based on their diagnostic status. Ultimately, the only notable shift in executive functioning was observed in the ASD and affective problems group, which demonstrated poorer outcomes.

## Introduction

Executive functions is an umbrella term that refers to a group of higher cognitive processes covering core functions such as planning, decision-making, error monitoring, emotion regulation, and behavioral inhibition (Doebel, 2020). Three main dimensions of executive functions commonly explored in cognitive assessment are inhibitory control, cognitive flexibility, and working memory (Doebel, 2020; Diamond, 2013). Inhibitory control refers to the cognitive ability to inhibit automatic responses or behaviors; deficits in this domain may lead to impulsive behaviors and social difficulties. Cognitive flexibility involves switching between thoughts or behaviors; deficits in cognitive flexibility may manifest as discomfort and difficulty with change (Munakata et al., 2011). Working memory is the ability to store and manipulate information temporarily; deficits in working memory may lead to difficulties in completing multi-step tasks and memory problems (Cowan, 2022).

From a neurodevelopmental perspective, executive function acquisition and maturation starts in early childhood, being a highly stable, heritable, and transdiagnostic trait-like feature of psychopathology (Friedman et al., 2016; Hatoum et al., 2018). Executive function deficits appear in varying degrees in child/adolescent psychopathology and have been associated with impairments in executive functioning, which in turn affects their ability to learn, self-care in daily life, and social behavior (Goschke, 2014; Duijkers et al., 2023). Executive functions approach in infancy is essential because their profile predicts a host of clinical outcomes in studies of adult psychopathology, including long-term functional recovery (Gruber et al., 2008), overall functioning (Martino et al., 2009), quality of life (Cotrena et al., 2016), and social/occupational functioning (O’Donnell et al., 2017).

The National Institute of Mental Health’s Research Domain Criteria (RDoC, NIMH) calls for the examination of transdiagnostic constructs across multiple levels of measurement (e.g., cognitive and domains of behavioral functioning) to improve the understanding of the underlying neurobiological underpinnings (Garvey et al., 2016). Regarding neuropsychological measures, Verbal Fluency Tasks (VFTs) are widely used in executive processing assessments globally (Lezak et al., 2012). VFTs are traditionally employed to assess verbal executive processing because they necessitate accessing lexical-semantic knowledge, switching between meanings, categories, or concepts, monitoring working memory representations, and inhibiting interfering thoughts (Amunts et al., 2020; Shao et al., 2014). Consequently, VFTs have been extensively researched as a potential screening tool for differentiating individuals within the context of various neuropsychiatric disorders (García-Herranz et al., 2020; McDonnell et al., 2020). Over the past two decades, Emotion Fluency Tasks (EFT) have emerged as a promising complementary modality to VFTs (Abeare et al., 2009). Previous studies have shown that EFT taps into similar cognitive processes as other verbal fluency tasks, suggesting its potential as a valid instrument for affective-cognitive measurement (Abeare et al., 2016; Suchy, 2011). In this vein, previous studies have demonstrated that performance on EFT was associated with sympathetic activation, such as heart rates, during task completion, whereas this phenomenon was unrelated to participants’ performance during non-affective fluency task completion (Abeare et al., 2016; Kennedy & Schley, 2000). These previous findings suggest that EFT may recruit broader and more diverse neural networks during performance compared to traditional verbal fluency tasks.

EF deficits are present during childhood in various neurodevelopmental disorders including both ASD and ADHD (Barkley, 1997; Ozonoff et al., 1999). Worldwide, 5.9%–7.1 % of children are currently diagnosed with attention-deficit/hyperactivity disorder (ADHD) [Polanczky et al., 2014]. The most common symptoms include inattention, hyperactivity/impulsivity, or combined. Children with ADHD often demonstrate executive dysfunction, which is believed to be responsible for many of their commonly observed behavioral problems (Wolraich et al., 2019). Autism Spectrum Disorder (ASD) is another neurodevelopmental disorder, with roughly 1 in 36 children identified as having ASD (Maennet et al., 2023). Children with ASD often exhibit impaired social interactions, limited communication skills, and stereotyped behavioral patterns (APA, 2022; Riddiford et al., 2022), These symptoms are usually seen before the age of three, which is also a critical period for children to develop cognition. Children with ASD typically have varying degrees of impairment in executive functioning, which in turn affects their ability to learn, self-care in daily life, and social behavior (Lee et al., 2023).

Children and adolescents who present affective symptoms also commonly show deficits in executive functions. This deficit can be state-independent and persist during affective episode remission (Lee, Hermens, Porter, & Redoblado-Hodge, 2012. In this psychopathological context, EF has received considerable interest as a potential endophenotype or critical underlying construct in affective psychopathology. Considering the transdiagnostic nature of executive deficits, this study aims to conduct an exploratory comparative analysis of data related to the neuropsychological assessment of executive functions and executive functioning in children and adolescents diagnosed with ADHD and ASD and who present affective symptoms. This study hypothesizes that neuropsychological markers help understand how executive deficits occur in each condition.

## Methods

### The Setting of Data Collection

Data for this cross-sectional study were collected from neuropsychological assessments conducted at the comprehensive outpatient specialized clinic in the Federal District, Brazil. The clinic provides medical, psychological, and speech therapy assessments for comprehensive diagnostic purposes. Referrals to the clinic come from various sources, including parents/patients, schools, pediatricians, psychologists, and psychiatrists. Participants voluntarily provided informed consent for all experiments, demonstrating their understanding and commitment to the study. This study was approved by the Federal University of Rio de Janeiro Ethics Committee (CAAE: 74921623.3.0000.5263), ensuring the ethical conduct of the research. Participants were excluded if they reported a history of genetic or neurological disorders.

### Behavioral and Affective Problems

The Strengths and Difficulties Questionnaire – SDQ (Goodman, 2001) assessed the parenting perception of social problems. The SDQ provides information about five domains: antisocial/conduct behaviors, peer relation problems, emotional issues, prosocial behaviors, and a measure of ADHD focusing on hyperactivity-inattention. Throughout the scale, parents were asked to rate their agreement on a three-point Likert scale ranging from 0: “not true” to 3: “certainly true,” allowing for a nuanced understanding of their perceptions. All variables from SDQ were considered for this analysis to characterize the sample included.

### IQ Scores

The Wechsler Intelligence Scales for Children, Fourth Edition (WISC-IV; Wechsler, 2008) and Adults, Third Edition (WAIS-III; Wechsler, 1991) were used to assess general intellectual development. The WISC-IV and WAIS-III provide full-scale IQ scores, which were transformed into age-normed scaled scores.

### Non-affective Executive Functions

To assess non-emotional verbal fluency abilities, the following tasks were administered to the participants: (1) Verbal fluency with semantic criteria (VFS): in this task, the participant must say as many words as possible that represent clothes or clothing in two minutes; (2) Verbal fluency with spelling criterion (VFSC): the order given to the participant is to say the most significant number of words that begin with the letter “p”, except proper nouns, in two minutes; (3) Free verbal fluency (FVF) requires the participant to name as many words as possible, except proper nouns and numbers, in two minutes and 30 seconds.

### Affective Executive Functions

The Emotional Fluency Task (EFT) is a measure of emotional word production and emotional vocabulary recall. Instructions are: “Give me as many emotion words as possible within one minute”. Two independent and blinded raters recorded and transcribed each participant’s responses for future classification. Verbal words such as “smiling” or adjectives such as “happy” were considered emotional terms and counted to generate the total number of words spoken. Furthermore, the words generated by the participant were categorized into primary emotions (happiness, sadness, disgust, anger, and fear) or secondary emotions (emotions that translate inferential meanings, such as longing and love). This classification considered the previous consensus among theorists about the five basic emotions (for a review, see Tracy & Randles, 2011). To categorize “complex emotions”, we considered the theory of constructed emotions, which considers that complex emotions result from acquired knowledge and has no categorization according to the independent nature of our perception of them.

The following scores are generated from EFT performance analysis: (1) total number of emotions spoken (NES); (2) number of perseverations (NP), i.e., the number of repeated responses); and (3) number of rule breaks (NRB), i.e., words that do not convey emotional meaning. They are divided into basic (joy, sadness, anger, disgust, and fear) or complex words. Terminological variations of the same meaning (e.g., “anger” and “angry” or “love” and “loving”) were considered as just one word. Unlike the other tests used in the study, the EFT did not have its basic psychometric properties tested in any previous study, which is why the analysis of construct validity and interrater reliability was conducted. Two clinical psychologists scored EFT independently to assess interrater reliability.

### Executive Functioning

The Behavior Rating Inventory of Executive Function-Second Edition (BRIEF-2), parent form (Gioia et al., 2015), is a parent-rated scale of executive functioning in children aged 5 to 18. It includes 63 multiple-choice items that are categorized into subscales of Inhibit, Shift, Emotional Control, Initiate, Working Memory, Plan/Organize, Organization of Materials, and Task Monitor. The Cronbach’s Alpha scores found in the Brazilian version are like those reported in the original version, indicating good internal consistency of the instrument in the three versions of BRIEF-2 (Carim et al., 2012). For the exploratory analysis, the T-scores of all subscales were considered.

## Data Analysis

Initially, data were screened using descriptive statistics. T-tests and correlations were conducted to assess whether age, sex, and IQ total scores were associated with performance on cognitive tasks. It examined correlations between variables to ensure that the assumption of no multicollinearity was met in regression analyses. The assumption of normality of the data was tested using Shapiro-Wilk’s test. Then, a one-way ANOVA or a MANOVA with Bonferroni’s post hoc was adopted to test the influence of the experimental condition on the tactical responses. This technique of including all dependent variables in the model was chosen to reduce the inflation of the type I error (Nobe, 2009). The level of significance was set at 5%. All analyses were conducted using the statistical software JASP Team (2024). JASP (Version 0.18.3) [Computer software].

In order to test the basic psychometric properties of EFT, the Interrater level of agreement was tested. Kappa values regarding the strength of level of agreement were classified following McHugh (2012): < 0.00 – poor; 0.00-0.20 – none; 0.21-0.39 – minimal; 0.40-0.59 – weak; 0.60-0.79 – moderate; 0.80-0.90 – strong; >0.90 – almost perfect. It used weighted Kappa due to the ordinal level of measurement of variables (Kottnet et al., 2011). Interrater reliability for the EFT was calculated with a Pearson’s correlation coefficient and intraclass coefficient for agreement. To examine construct validity, correlations between the ETF and verbal fluency task (VFT), phonemic verbal fluency - PVF (p phonemic), and semantic verbal fluency -SVF (clothes) were calculated to examine construct validity.

## Results

### Sample characteristics

A hundred thirty-four children and adolescents, with a gender distribution of 57.4% males, were enrolled in this analysis. The participants were aged between 7 and 17 years old (mean=11.7, SD=2.6), were enrolled in this analysis. Sixty-two (46.2%) participants were included in ADHD group, forty-one (30.5%) were included in ASD group, and 31 (23.1%) participants were diagnosed with high levels of anxious-depressive symptoms.

A one-way ANOVA did not show a significant difference between parents’ reports regarding pro-social behaviors [F(2,117)=.868, p=.422], hyperactivity symptoms [F(2,117)=.700, p=.499], emotional problems [F(2,117)=2.381, p=.097] or antisocial behaviors [F(2,117)=1.380, p=.256] when compared three groups. However, a one-way ANOVA adjusted by Bonferroni’s method showed a significant difference regarding parents’ reports about social relationships between the three groups [F(2,117)=7.766, p<.001], and a posthoc analysis showed a significantly higher social isolation index in ASD and Affective Disorders groups when compared with ADHD group.

**Table 1.**
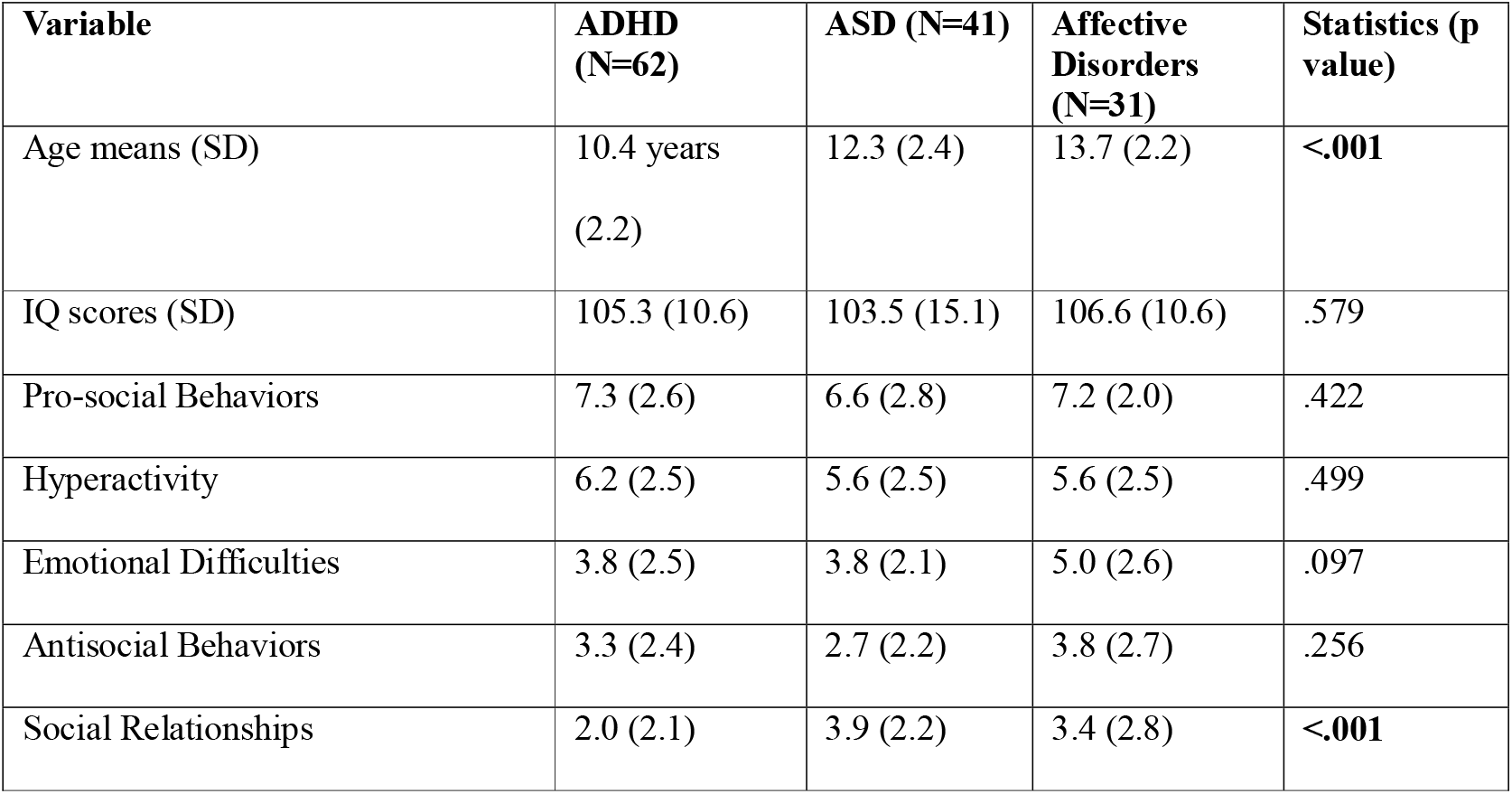
Sample characteristics and dimensional assessment of problems according to group.

### Executive Functions’ Tasks

#### I Non-affective executive functions

A Pillai’s MANOVA showed significant differences between the three groups regarding the performance in FVF [F(121,2)=4.457, p<.05]; VFS [F(121,2)=8.158, p<.001]; and VFSC [F(121,2)=6.234, p<.01]. A Bonferroni’s adjusted post hoc analysis revealed a significant difference between the affective disorders group and other groups in all VFT (t=-2.878, p<.05), VFS (t=-4.02, p<.001), and VFSC (t=-3.448, p<.01), but no between ASD and ADHD groups.

#### II Affective executive functions

##### a) Basic Psychometric Properties of EFT

Interrater level of agreement was classified as almost perfect for NES [k = .961 (IC 95% .889-1.0)], NP [k = .988 (IC 95% .966-1.0), and strong for NRB [k = .828 (IC 95% .685-.972). Intraclass correlation for agreement for NES was rICC = 0.99 (IC 95% .995-.998); for NP was rICC = 0.988 (IC 95% .982-.992); and for NRB was rICC = 0.834 (IC 95% .764-.884). Regarding construct validity, Pearson’s correlations between NES and VFT, PVF, and SVF were analyzed. The number of emotions produced in the EFT was correlated with the number of words produced in VFT (r=.632, p<.001); PVF (r=.602, p<.001); and SVF (r=.552, p<.001). These results provided convergent validity between EFT and other verbal fluency measures.

##### b) Comparative Groups’ Performances

A one-way ANOVA did not show a significant difference between the three groups compared to the total number of words generated in EFT [F(131,2)=1.003, p=.369]. Further, no significant differences were found between the three groups when considering the perseverance and broken rules indexes and the number of complex emotions generated. See Table 2 for descriptive statistics about the performance of groups.

**Table 2.** Performances in Emotional Fluency Task according to group.

| **EFT variables** | **ADHD** | **ASD** | **Affective**<br>**Disorders** | **Statistics** |
| --- | --- | --- | --- | --- |
| Total scores | 8.323(2.65) | 8.829 (3.72) | 9.258 (2.97) | F(123,2)=1.003,<br>p=.369 |
| Perseverations | .310 (.706) | .300 (.687) | .357 (.678) | F(123,2)=.061,<br>p=.940 |
| Rule Breaks | .241 (.601) | .250 (.543) | .143 (.067) | F(123,2)=.395,<br>p=.674. |

It was found a significant correlation between EFT total scores and IQ scores (r=.279, p<.001). A linear regression analysis suggested QI scores as a predictor of EFT performance [F(130,1)=10.856, p<.001].

#### Parents’ Reposts about Executive Functioning

In order to avoid the homogeneity of covariance between dependent variables, a one-way ANOVA was conducted to compare the BRIEF-2 variables between the three groups. Regarding the subdomains of executive functioning analysis through parents’ report, only shift subdomain [F(98,2)=3.756, p<.05] appeared as a significant difference between the three groups. A Turkey Post hoc analysis did not reveal a significant difference between ASD and Affective Disorders groups (t=1.86, p=.155). Still, it showed a significant difference when compared with the ADHD group (t=-2.7, p<.05). The statistics of comparison between the three groups regarding executive functioning variables are shown in the table below.

**Table 3.** Executive Functioning indexes comparisons between the three groups.

| **EF Domain** | **ADHD** | **ASD** | **Affective**<br>**Disorders** | **Statistics** |
| --- | --- | --- | --- | --- |
| Inhibit | 59.5 (13.1) | 60.8 (11.5) | 62.8 (14.7) | P=.618 |
| Shift | 62.8 (11.0) | 70.4 (12.4) | 64.1 (13.1) | **P<.05** |
| Emotional Control | 59.3 (12.3) | 58.7 (11.0) | 63.6 (12.1) | P=.302 |
| Initiate | 59.6 (6.6) | 62.4 (11.9) | 62.0 (11.6) | P=.401 |
| Plan/Organize | 60.6 (11.9) | 60.1 (10.9) | 62.9 (14.2) | P=.758 |
| Working Memory | 66.9 (7.8) | 64.4 (14.2) | 61.0 (13.8) | P=.124 |
| Task Monitor | 61.2 (9.1) | 61.2 (13.0) | 62.4 (11.5) | P=.90 |
| Organization of Materials | 62.3 (10.1) | 60.7 (10.6) | 59.1 (11.6) | P=.481 |

## Discussion

This study focuses on direct comparisons of both cognitive and functioning levels of executive functions in children and adolescents with ADHD, ASD, and Affective Problems. It represents a deeper investigation of deficits related to executive functions as a transdiagnostic manifestation by including measures of affective and non-affective executive functions. The emotional fluency test, used to investigate affective executive functions, demonstrated favorable psychometric properties.

The results of this study align with previous research, as they found no evidence to support the hypothesis that children and adolescents with ASD and those with ADHD have distinct affective and non-affective executive functions cognitive profiles. This result is consistent with a previous metanalytic study conducted by Townes et al. (2023) that did not show a significant difference between the comparison of executive functions profile in children and adolescents with ASD and those with ADHD. However, this meta-analysis did not include measures focused on assessing affective executive functions. Carrick & Hamilton (2023) shown in a previous study focusing on comparison of affective executive functions in children with ADHD and with autistic-traits and did not find a significant differences according to diagnostic state. However, it is worth highlighting that this study used a different methodology to measure affective executive functions.

Regarding executive functioning data, this study showed a significant difference between ADHD and ASD when compared to shift indexes. However, the results did not show a significant difference between ASD and affective disorders groups, which indicates that both ASD and affective disorders groups present deficiencies in behavioral flexibility. Flexibility is typically thought to comprise both cognitive and behavioral components. ‘Cognitive flexibility’ is broadly defined as the mental ability to switch between thinking about two different concepts according to the context of a situation, and ‘Behavioural flexibility’ refers to the adaptive change of behavior in response to changing repetitive behaviors (RRBs), considered core deficits in ASD, can include stereotyped movements, insistence on sameness, and circumscribed or perseverative interests (APA, 2022). The severity of RRBs is associated with measures of cognitive inflexibility in ASD (Lopez et al., 2005). Studies of the neural circuitry underlying RRBs transdiagostically point to a critical role for frontostriatal systems in mediating these behaviors (Wilkes et al., 2018). A recent review of neural mechanisms underlying cognitive and behavioral flexibility in autism also points to atypical lateral-frontoparietal Network patterns and midcingulo-insular Network activation in response to task switching and set-shifting (Uddin, 2021).

On the other hand, high levels of affective symptoms involve excessive worry or the tendency to dwell on difficulties and perceive future problems as more likely than they are in real situations. In contrast, depression involves rumination or passively focusing on distressing thoughts in response to sad moods and experiences (APA, 2022). Worry and rumination may reflect the same underlying construct of repetitive negative thinking, which is likely a product of inflexible thinking and difficulty engaging the lateral frontoparietal Network executive control systems in the service of emotion regulation (Burrows et al., 2017). Based on this theoretical assumption, the results of this study suggest that data related to cognitive flexibility, but not to behavioral flexibility, serve to differentiate children and adolescents diagnosed with ASD from those diagnosed with affective problems.

Almost all the DSM-5 categories include clinical conditions in which domains of executive functioning are compromised. This raises the question of how much further analysis of executive functions and executive functioning has discriminative value. Results extracted from this analysis reinforce the purpose of The National Institute of Mental Health’s Research Domain Criteria (RDoC, NIMH), which highlights the need to examine executive dysfunctions using methods that assess multiple domains of functioning to assist with fully understanding transdiagnostic impairments. Multimethod findings can support initial steps for improving experimental procedures and intervention regimens to promote executive dysfunctions improvement, along with further refining the conceptualization of the nature of the deficits.

A strength of the current study was the comparative analysis regarding executive dysfunctions presented by children and adolescents in a dimensional perspective and a multilevel analysis, including affective executive functions. Regarding the limitations of this study, it was not examined how some clinical dimensions might moderate associations between neurodevelopmental symptoms severity and EF. Previous research has indicated that comorbid additional clinical symptoms are associated with more severe cognitive problems, such as oppositional-defiant symptoms, which may exacerbate executive function difficulties in neurodevelopmental disorders (Block et al., 2022). In the same way, although some comorbidities might have enhancing moderating effects because they are associated with greater executive dysfunctions, anxiety symptoms may mask specific EF difficulties in neurodevelopmental disorders (Anning et al., 2023); however, examining these complex interactive and potential counteracting executive functions mechanisms was beyond the scope of our study. Finally, this analysis did not include direct comparisons, allowing for tighter control over the tasks, task conditions, and factors such as age, sex, and medication status, which could confound the interpretation of results of indirect comparisons where participants’ data are first compared to typically developing individuals.

## Data Availability

All data produced in the present work are contained in the manuscript

## Acknowledgments

We thank the anonymous reviewers for their insightful comments.

